# Confounders in the relationship between temporomandibular disorders and posture: a scoping review protocol

**DOI:** 10.1101/2023.05.16.23288058

**Authors:** Pier Claudio Diciolla, Paolo Bizzarri

## Abstract

**Background:** The relationship between temporomandibular disorders (TMDs) and posture is widely discussed in literature. However, evidence to support this relationship is scarce, and little is known about their causal association.

**Objectives:** The aim of this scoping review is to provide an analysis on how much the potential confounders (craniofacial morphological elements, sleep apnea and/or psychological factors) are the real responsible for postural changes.

**Methods:** A search will be conducted through all publications written in English and Italian about this topic using Mediline (PubMed) database. Cross-sectional studies will be included concerning morphological (e.g. class II, class III, crossbite, etc.), respiratory (e.g. sleep apnea) and/or psychological (e.g. anxiety, depression) elements and postural patterns (lordosis, kyphosis, scoliosis, forward head position) in adults (≥18 years).

No restriction for the year of publication.

The level of bias in the included studies will be assessed using the JBI Critical Appraisal Checklist for Analytical Cross-Sectional Studies.

## INTRODUCTION

### Background

Temporomandibular disorders (TMDs) are defined as a set of musculoskeletal and neuromuscular conditions that involve the temporomandibular joint (TMJ), the masticatory muscles, and all associated structures. Pain associated with TMDs is clinically expressed as muscle and/or joint pain, aggravated by chewing and other jaw activities. It may be associated (but not necessarily) with TMJ biomechanical dysfunctions (TMJ clicking or locking and jaw movement limitations). Chronic forms of temporomandibular pain can lead to limitations in work activity or social interactions, resulting in an overall reduction of quality of life. (1)

The reference classification for TMD is the *Diagnostic Criteria for Temporomandibular Disorders (DC/TMD)* (2).

The relationship between TMDs and posture is widely discussed in literature with most studies rejecting a clinical association (3-10).

However, there are regional factors that may mimic a relationship between orofacial pain, occlusion and specific postural features. In particular, obstructive sleep apnea is associated with assuming a forward and extended head position to improve airway patency and maintain the visual sense (11); similarly, studies support a morphological relationship between occlusal factors and head position, as subjects with class II and hypodivergent pattern seem to have a more forward and extended head, while subjects in class III tend to have a more flexed head (12,13,14); finally, psychological factors such as anxiety, depression and stress also seem to play a role in assuming different postural patterns (15,16).

Recent reviews were found on spinal postural patterns regarding obstructive sleep apnea (11), or occlusal factors (13,17,18). Therefore, there is currently no review in the literature that takes into account the various confounders that can mimic a correlation between TMD and posture.

It’s necessary to carry out a scoping review on the subject to find sufficient evidence to accept or reject this relationship and to guide future research.

### Objectives

The aim of this scoping review is to provide an analysis on the association of specific orofacial factors (occlusion, craniofacial morphology, sleep apnea and/or psychological factors) with specific postural patterns.

## METHODS

The results of this scoping review were reported using the Preferred Reporting Items for Systematic reviews and Meta-Analyses (PRISMA 2020) extension for Scoping Reviews (PRISMA-ScR) Checklist (19).

### Elegibility Criteria

Only cross-sectional studies with adult participants (≥18 years) were included in this review. Studies that accepted underage participants, patients with serious pathologies (neurological diseases, cancer, rheumatoid arthritis, traumas, etc.) were excluded.

Studies in Italian and English languages were selected, from any year of publication, concerning craniofacial occlusal or morphological (e.g. class II, class III, crossbite, etc.), respiratory (e.g. sleep apnea) or psychological (e.g. anxiety, depression) elements. Outcome measure is represented by the presence of differences in the postural pattern (e.g. lordosis, kyphosis; scoliosis; forward head position).

### Information sources

The searches were conducted on the PubMed database (Medline), started on February 7, 2023, and ended on February 21, 2023.

### Search

The search strategy was designed according to the specific settings of the reference database (PubMed). The search strategy was developed according to the PICOS model of the research query: free terms or synonyms were used, in combination with the Boolean operators (AND, OR); no filters were applied by year of publication.

Furthermore, a manual search was conducted through the bibliographies of the results reviewed in order to implement additional studies eligible for those included through the initial screening.

The following search strategy was used:

*((Posture or scoliosis or kyphosis or lordosis or alignment or “Forward Head” or “head tilt”) AND (cervical or neck or thoracic)) AND (“sleep apnea” or OSAS or malocclusion or “class II” or “class III” or crossbite or hypodivergent or hyperdivergent or cephalometric or morphometric or mood or anxiety or depression or psychological)*

### Selection of sources of evidence

The process for selection sources of evidence was performed individually by a single reviewer (PCD), under the supervision of a second author (PB).

The research results were managed with the “EndNote 20” reference management software (20), where the records were imported, allowing the identification and elimination of duplicates and, subsequently, the screening of titles and abstracts.

Subsequently, the *full-texts* of the identified studies were searched for further evaluation according to the pre-set eligibility criteria.

### Data charting process

A data extraction form (based on the form used by Li et al., 2015 (21,22)) was developed, tested on three of the included studies, and refined accordingly. Data were collected by a single reviewer (PCD) through *full-text* reading of included studies, supervised by a second author (PB).

### Data items

For each article the following were extracted:

- *first author’s surname and year of publication;*
- *article’s title and journal of publication;*
- *objective of the study;*
- *population (sample size, demographic characteristics of the sample);*
- *diagnostic method*
- *postural outcome (domains assessed, evaluation criteria used);*
- *results of the study*
- *conclusions of the study*.

### Critical appraisal of individual sources of evidence

The *JBI Critical Appraisal Checklist for Analytical Cross Sectional Studies* (23), developed by the Joanna Briggs Institute (2017), was used to assess the risk of bias in each of the studies included in the review.

This checklist is made up of eight specific items:

1. *Were the criteria for inclusion in the sample clearly defined*?
2. *Were the study subjects and the setting described in detail?*
3. *Was the exposure measured in a valid and reliable way?*
4. *Were objective, standard criteria used for measurement of the condition?*
5. *Were confounding factors identified?*
6. *Were strategies to deal with confounding factors stated?*
7. *Were the outcomes measured in a valid and reliable way?*
8. *Was appropriate statistical analysis used?*

For each question, the possibilities for answering are as follows: “*Yes*”, “*No*”, “*Unclear*”, “*Not applicable*”.

The evaluation was performed by a single reviewer (PCD)

### Synthesis of results

Due to the heterogeneity of outcome measures and participants, it was not possible to pool together the results from the included studies in a statistical form; therefore, it was decided to report them in narrative form.

This manuscript has been seen and approved by all listed authors.

The authors have declared no competing interest.

## Data Availability

All data produced in the present study are available upon reasonable request to the authors.

## Notes

### Funding Statement

This study did not receive any funding.

### Author Declarations

The source data were openly available before the initiation of the study (https://pubmed.ncbi.nlm.nih.gov).

